# Repetition suppression of motor cortex may predict responsiveness to high-frequency rTMS in chronic pain

**DOI:** 10.1101/2025.08.12.25332182

**Authors:** Shohreh Kariminezhad, Selja Vaalto, Laura Säisänen, Mervi Könönen, Erika Kirveskari, Vilma Mannila, Jelena Hyppönen, Jarmo Laine, Jari Karhu, Petro Julkunen

## Abstract

**Background:** High-frequency repetitive transcranial magnetic stimulation (rTMS) has been reported to yield promising, albeit variable, outcomes in chronic pain therapy and management. A factor contributing to the treatment success is individual neuroplastic capacity. Neuroplasticity can be assessed in various ways with TMS, *e.g.* through neural repetition suppression (RS) based on habituation. The present study aimed to evaluate the TMS-induced RS in patients with chronic pain, undergoing conventional rTMS treatment, and observing whether RS is indicative of the experienced analgesic effects.

**Methods:** Twenty-one patients received standard 10-Hz rTMS treatment in five or ten daily sessions. Twenty RS trials, each consisting of four TMS pulses given 1 s apart, were given and assessed for motor evoked potentials (MEPs) before the first rTMS therapy session. For analysis purpose, the RS paradigm was evaluated through two states: 1) “*dynamic*” state (spontaneous excitability), first to second pulse, and 2) “*stable*” state (suppressed excitability), suppressed responses induced by second to fourth pulse. Analgesic effect from rTMS was assessed using Brief Pain Inventory (BPI), and painDETECT screening questionnaire, both conducted before and after the rTMS treatment.

**Results:** Both the dynamic and the stable state RS exhibited good accuracy (0.789 – 0.944) in identifying patients showing analgesic effect in response to rTMS. Also, significantly greater suppression of MEPs (*p*<0.05) were observed in the RS between those who benefitted from rTMS and those who did not.

**Conclusions:** These indicative findings provide support for the potential of RS to set a premise for improved individualized neuromodulation treatments.

## 1 Introduction

Acute pain serves as a protective mechanism through which the body triggers adaptive withdrawal responses to noxious or potentially noxious stimuli. The mechanisms underpinning the pain processing encompasses a well-defined brain network composed primarily of somatosensory cortex and thalamus (Apkarian et al., 2005). However, the maladaptive mechanisms may result in recurrent or persistent pain beyond normal healing time, referred to as chronic pain. Pain is considered as chronic when it persists for at least 3 months (Treede et al., 2015). Despite the substantial advances in analgesic interventions, the treatment success rate is low (Cruccu et al., 2016; Dansie & Turk, 2013; Finnerup et al., 2010; Säisänen et al., 2022; Wiffen et al., 2013).

Repetitive transcranial magnetic stimulation (rTMS) is considered as a non-invasive neuromodulation intervention, administrated in a subset of chronic pain patients in whom the conventional therapies have failed (Lefaucheur, 2006; Lefaucheur et al., 2001). Although the underlying mechanisms remain elusive, high-frequency rTMS, applied over M1, has been demonstrated to yield the greatest, albeit variable, analgesic effect (Lefaucheur et al., 2020; O’Connell et al., 2018). Multiple factors underpin the inconsistent treatment outcomes among which neuroplasticity might act as a key contributor. Neuroplasticity can determine the degree to which the neural pathways at cortical and subcortical levels are recruited, *e.g.* while recovering from a stroke or brain injury (Bavelier & Neville, 2002; Pascual-Leone et al., 2005). Yet, maladaptive neuroplasticity can be the core pathology in developing neuropathic pain, *e.g.* following spinal cord injury (Brown & Weaver, 2012).

Neuroplasticity has been demonstrated to associate with repetition suppression (RS), *i.e.,* an inherent brain mechanism indexed by attenuated neural activity when exposed to an identical stimulus (Grill-Spector et al., 2006; Julkunen et al., 2018). In the motor cortex, RS has been quantified with TMS, reflected as decrement of motor evoked potential (MEP) (Löfberg et al., 2014; Löfberg et al., 2013). TMS-induced RS has been suggested to reflect the interplay of two states: *dynamic* and *stable* (Kariminezhad et al., 2020). Dynamic state RS refers to the initial decrease of MEP amplitude in response to an intense and novel stimulus and is hypothesized to be indicative of efficient processing of the stimulus. However, the stable state of RS, that is the suppressed excitability observed by the diminished MEP amplitudes from the second to the fourth (last) stimulus, can be decomposed and assessed for two components itself. First component, *i.e.* the TMS-evoked response average over the suppressed responses, can be viewed as the overall neural excitability that is less influenced by the ongoing oscillatory dynamics. However, the second component, the changes from the second response to the last one, potentially imply the existence of an automatic memory through which the recent exposed stimulus has been retained on a timescale of seconds (Kariminezhad et al., 2020; Pitkänen et al., 2017). It has been shown in healthy subjects that the MEP amplitudes of this component of the stable state tend to remain within a given range with low variance following induction of short-term plasticity (Kariminezhad et al., 2020).

In the present observational study, our objective was to evaluate the potential of RS for predicting the individual rTMS responsiveness to conventional rTMS therapy in heterogeneous group of patients with chronic pain and observing whether RS is indicative of the experienced analgesic effects.

## 2 Methods

### 2.1 Subjects

Twenty-one patients (14 females, age range: 25-87 years, mean ± SD: 59 ± 17 years) with chronic severe drug-resistant neuropathic or assumed neuropathic pain or complex regional pain syndrome (CRPS) type I or II were included in this study, that was conducted in Kuopio University Hospital and Helsinki University Hospital (during years 2019–2020).

Demographic information of the patients is presented in Table 1. Patients filled the Brief Pain Inventory (BPI) (Cleeland & Ryan, 1994; Daut et al., 1983; Tan et al., 2004) and painDETECT (Freynhagen et al., 2006) questionnaires prior to the first and after the last treatment sessions. All patients provided a written informed consent prior to their participation. The study was approved by the research ethics committee of the Kuopio University Hospital (949/2018).

**Table 1.** Description of patients and pain-related information with individual rMT.

| No. | Sex | Pain |  | rMT | painDETECT |  |  | BPI |  |  | Therapy duration (days) |
| --- | --- | --- | --- | --- | --- | --- | --- | --- | --- | --- | --- |
|  |  | Topography | Etiology |  | Initial intensity score (NRS) | Change in intensity score | Change in total score | Initial severity score | Change in severity score | Change in impact score |  |
| 1 | F | face | atypical, iatrogenic | 36 | 2 | 0 | 1 | 4.50 | -0.25 | -3.71 | 5 |
| 2 | M | face | atypical, post-traumatic | 35 | 3 | 0 | -2 | 4.00 | -0.50 | -0.71 | 10 |
| 3 | F | face | atypical, iatrogenic | 44 | N.A. | N.A. | N.A. | 8.50 | -0.75 | -0.86 | 10 |
| 4 | F | face | atypical, iatrogenic | 34 | 3 | 2 | -4 | 4.25 | 0.00 | -1.86 | 5 |
| 5 | F | face | atypical, iatrogenic | 66 | 7 | -2 | -3 | 6.75 | 0.00 | -1.86 | 5 |
| 6 | F | face | atypical, iatrogenic | 33 | 5 | -3 | 1 | 3.50 | -1.25 | -1.57 | 5 |
| 7 | F | face | atypical, idiopathic | 44 | 2 | -1 | -9 | 2.00 | -0.75 | -1.86 | 10 |
| 8 | F | face | atypical, iatrogenic | 42 | 6 | -2 | -5 | 7.25 | -2.00 | 0.43 | 5 |
| 9 | F | hand | CPSP | 50 | 10 | 0 | 4 | 5.50 | 1.25 | 0.14 | 9 |
| 10 | F | face | atypical, post-traumatic | 48 | 3 | 1 | -2 | 4.50 | -0.36 | -1.57 | 10 |
| 11 | M | face | atypical, idiopathic | 29 | 7 | 0 | 7 | 7.27 | -0.52 | -0.14 | 10 |
| 12 | M | leg | CPSP | 40 | 5 | -1 | 0 | 2.45 | 1.80 | 0.14 | 10 |
| 13 | F | face | atypical | 46 | N.A. | N.A. | N.A. | 5.73 | -2.98 | -3.86 | 10 |
| 14 | F | face | trigeminal neuralgia | 38 | 7 | 0 | 0 | 5.27 | 1.73 | 1.00 | 10 |
| 15 | M | face | postherpetic neuralgia | 46 | 8 | 0 | 0 | 6.36 | 0.64 | 0.00 | 10 |
| 16 | F | face | atypical | 35 | 5 | -3 | -5 | 4.18 | N.A. | N.A. | 10 |
| 17 | F | face | trigeminal neuropathy | 33 | 5 | 1 | 3 | 3.18 | 1.18 | 1.29 | 10 |
| 18 | F | upper limb | CRPS | 78 | 7 | -1 | 1 | 7.09 | -1.34 | -2.57 | 10 |
| 19 | M | upper limb | CPSP | 48 | 7 | 0 | 16 | 7.00 | -2.00 | -2.00 | 10 |
| 20 | M | leg | CRPS | 29 | N.A. | N.A. | N.A. | 8.45 | N.A. | N.A. | 10 |
| 21 | M | face | N.A. | 39 | 6 | -2 | -2 | 6.45 | -1.95 | -0.86 | 10 |

BPI: Brief Pain Inventory. CRPS: Complex regional pain syndrome. CPSP: Central post-stroke pain. NRS:
Numeric rating scale. N.A.: Data not available.

### 2.2 Transcranial Magnetic Stimulation

For individual MRI-guided navigated TMS (nTMS), structural T1-weighted magnetic resonance images (MRIs) were acquired with a 1.5 T or 3T clinical MRI scanner (Philips Achieva, Philips, Eindhoven, The Netherlands; GE Signa, GE Healthcare, Chicago, IL, USA; or Siemens Avanto, Skyra and Aera, Erlangen, Germany) prior to the treatment. TMS was performed using navigated TMS (NBS System 4.3 or NBS System 5, Nexstim Oyj, Helsinki, Finland) with an air-cooled figure-of-eight coil and biphasic stimulation. The stimulation procedure was initiated by determining the cortical abductor pollicis brevis (APB) representation where the maximal amplitudes of MEPs were elicited, referred to as APB “hotspot” (Julkunen et al., 2009). Once the hotspot was located, coil rotation angle was optimized by maximizing the MEP amplitude and selecting the rotation angle that produced consistently highest amplitudes. The resting motor threshold (rMT) was then assessed using a system-integrated iterative threshold assessment tool with inter-trial interval of at least 5 s (Awiszus & Borckardt, 2012; Julkunen et al., 2012; Kallioniemi et al., 2018). Thereafter, twenty trains of four single TMS stimuli with an inter-stimulus-interval (ISI) of 1 s during the stimulus trains and an inter-train interval (ITI) of 17 s were employed at 110% rMT in RS paradigm. The paradigm, lasting 6 minutes, was employed prior to the first rTMS session (Fig. 1). The resulting TMS-induced MEPs were recorded using an integrated electromyography (EMG) system (Nexstim Oyj) at a sampling frequency of 3 kHz. A pair of Ag-Cl surface electrodes was utilized with the active electrode overlying the right APB muscle. The recorded data were subsequently processed offline in MATLAB (R2018b, MathWorks Inc., Natick, MA, USA). The MEPs with amplitude lower than 50 µV were regarded as non-responses.

**Figure 1.**
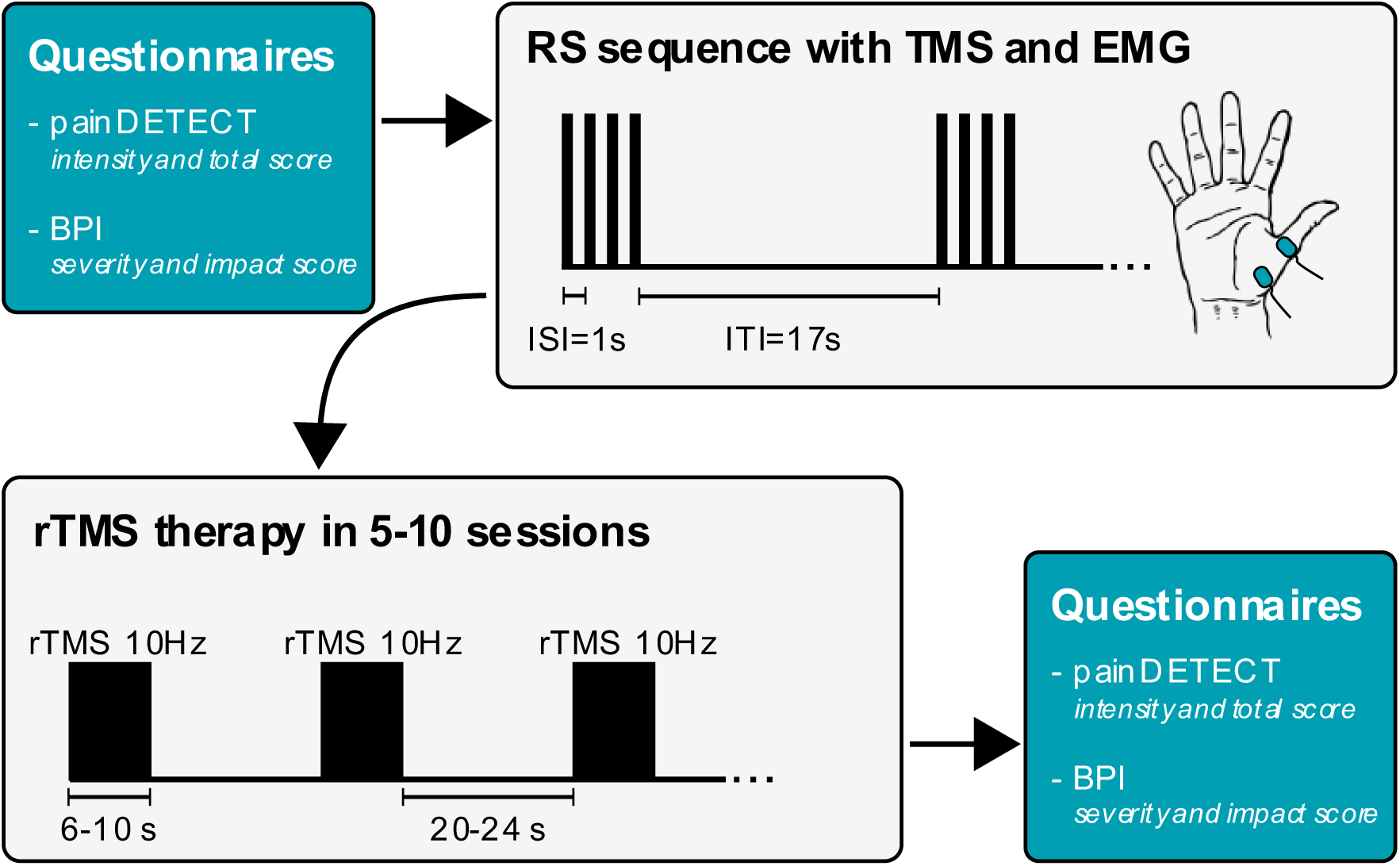
RS paradigm employed and assessed for MEPs prior to the first rTMS intervention. Patients received 5 or 10 sessions of 10-Hz rTMS. Two slightly different 10-Hz rTMS protocols were employed over the representative motor cortical area of the affected site, in two centers where the study was conducted. To evaluate the impact of rTMS on the pain severity, self-report questionnaires were completed prior to the first and the last sessions.

### 2.3 Repetitive transcranial magnetic stimulation treatment

Patients received 10-Hz rTMS treatment within either 5 or 10 consecutive weekdays over the representative primary motor cortex corresponding to the pain affected site. Due to non-responsiveness, seven patients received the treatment over secondary somatosensory cortex (S2) contralateral to the pain side following the fifth session. The number of sessions was decided based on whether the patient received treatment for the first time (10 days) or had had prior treatment (5 days). Accordingly, twelve patients received 10 sessions, and eight patients received 5 sessions of rTMS. One patient underwent 9 sessions of rTMS treatment missing one day of therapy. Two slightly different protocols were administrated in each center, according to their standard protocols. The patients in Kuopio received 10-Hz rTMS with trains of 6 s and ITI of 24 s (for a total number of 3,000 stimuli during each session).

The patients in Helsinki received the 10-Hz rTMS treatment with trains of 10 s and ITI of 20 s (for a total number of 3,030 stimuli during each session).

### 2.4 Treatment outcome after rTMS

The baseline condition prior to the first and outcome prior to last rTMS sessions was rated using the BPI questionnaire, and the painDETECT. BPI questionnaire consists of five items, assessing two dimensions of pain: severity and impact. First four items are used to rate the *pain intensity* on an 11-point scale (0 = no pain to 10 = worst pain ever). The mean score from the four pain intensity items represents the *pain severity*. The last item of the BPI consists of 7 sub-items from which their mean score is used to evaluate the extent to which the pain has *interfered with seven daily activities* such as walking, enjoyment of life, and mood (impact score). Higher scores indicate more interference. For painDETECT, the score on nine-item version of questionnaire consisting of seven sensory symptom items, one pain- course pattern item, and one item on pain radiation was calculated (ranging 0–38) (Cappelleri et al., 2014; Freynhagen et al., 2006). In addition, numeric rating scale (NRS, ranging 0–10) to estimate pain intensity at the moment of filling the questionnaire was included in the painDETECT questionnaire. To determine the analgesic effect to rTMS intervention, the absolute change in the scores on painDETECT and BPI were evaluated between the first and last session of treatment, including the BPI severity score, BPI impact score, and painDETECT intensity and total scores. Three subjects did not complete the painDETECT questionnaire at baseline and two did not complete BPI questionnaire at follow-up.

### 2.5 Analysis

MEP-data were first averaged over all the trains on the basis of their ordinal position within a single train, per subject. The dynamic state of RS was assessed by dividing the mean MEP amplitude induced with the second stimuli in the RS sequence by the mean MEP amplitude induced with the first stimuli in the RS sequence. The stable state of RS, *i.e.* suppressed level response was evaluated via calculating the mean of the second, third, and fourth MEP amplitudes in the RS sequence (Fig. 2). The change in stable state of RS was defined as the mean of the differences between the consecutive MEPs from the second to fourth responses (mean of the difference between the second stimulus and the third one, and between the third and the fourth one) indicating recovery rate towards non-suppressed state.

**Figure 2.**
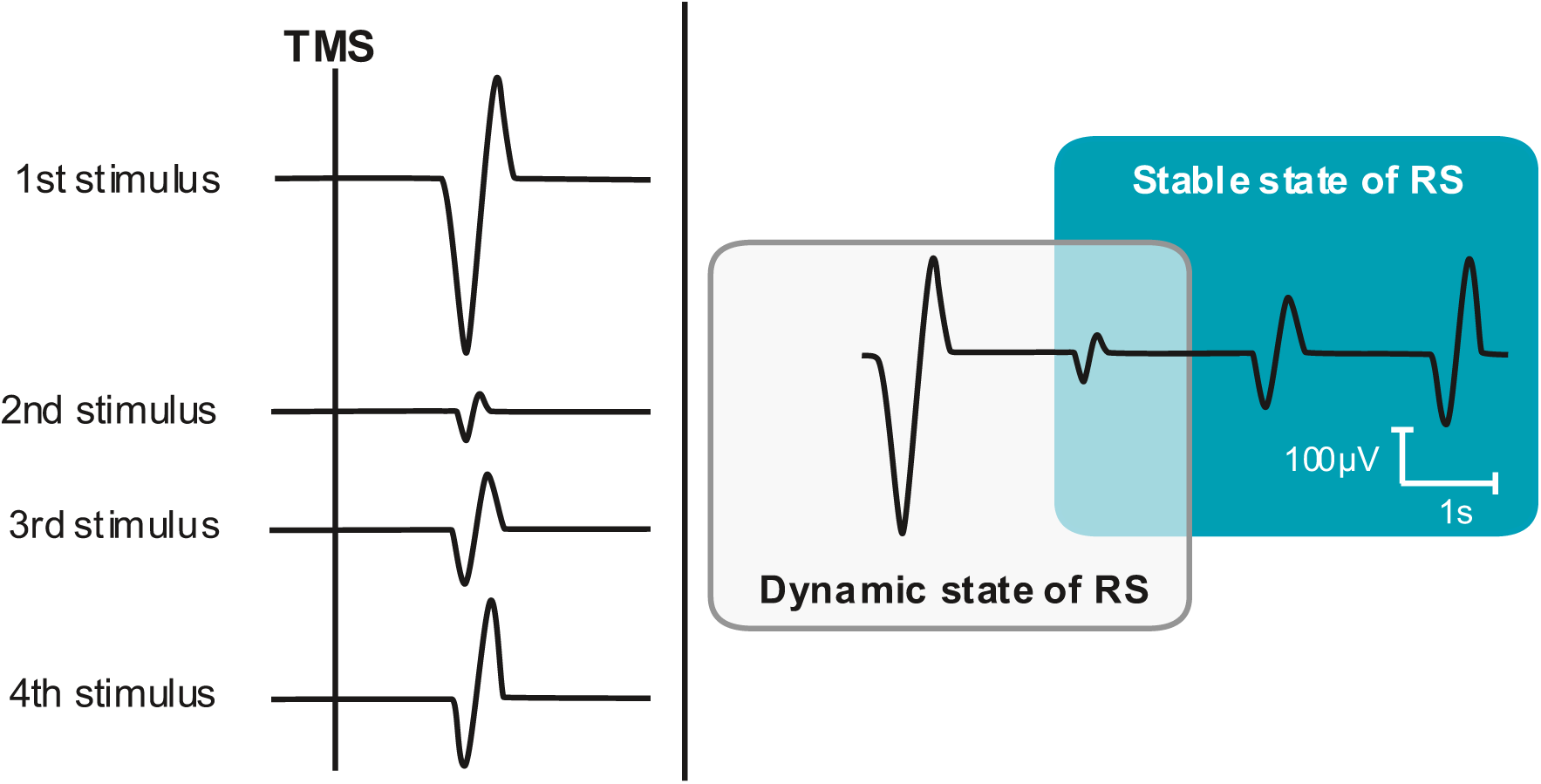
A schematic example of TMS-induced repetition suppression (RS). RS has been hypothesized to reflect the interplay of two states: *dynamic* and *stable*. For assessment purpose, the dynamic state has been depicted as the ratio of the average second MEP amplitude to the average first MEP amplitude, whereas the stable state was evaluated from two aspects: 1) the overall excitability of the stable state was calculated as the average MEP amplitude over the second, third, and fourth MEPs, and 2) the change in stable state of the RS was calculated as the average of the differences between consecutive responses.

For the statistical measures of the discriminative ability of the dynamic state of RS, stable state of the RS and changes in stable state of RS, area under the curve (AUC) and accuracy analysis of receiver operating characteristic (ROC) curve were applied. Those likely/not likely to benefit from the rTMS treatment were defined for each assessed RS response and outcome score by optimizing the cut-off value for the outcome scores to maximize the AUC. The minimum number of patients included within each group was set to five. The optimization was performed running ROC analysis with all possible score values between minimum and maximum score and choosing the score value for discriminating threshold resulting in greatest AUC value while considering the set constraint of the group sizes. The cut-off values for the assessed RS responses were determined from the optimized ROC curves looking for the point that is closest to the upper left corner in the ROC curve. Comparison of the RS-parameter values between the classified patient groups were performed using Mann-Whitney U test.

The statistical analysis was conducted using SPSS (v. 25.0, SPSS Inc., IBM Company, Armonk, NY, USA) and MATLAB (version R2021b, MathWorks Inc., Natick, MA, USA), and *p* < 0.05 indicated statistical significance.

## 3 Results

The optimal cut-off points for the ROC curve analyses for each tested parameter are shown in Table 2. Below, we summarize the findings that demonstrate most potential based on the classification accuracy demonstrated in Table 2.

**Table 2.** AUCs and optimal cut-off points for patients likely to benefit/unlikely to benefit from the 10-Hz rTMS based on the parameters of interest. The number of patients who showed benefit vs. those who showed no benefit based on the optimal cut-off points for changes in assessed scores. Asterisk (*) indicates statistically significant difference in the parameter values between the patients likely and unlikely to benefit (*p*<0.05, Mann-Whitney U tests).

| **Outcome scores and assessed parameter** | **Accuracy (AUC)** | **Optimal cut-off for change in score** | **Number of subjects likely to benefit/unlikely to benefit** |
| --- | --- | --- | --- |
| **BPI severity score** |  |  |  |
| Dynamic state of RS | **0.895 (0.833)\*** | -1.2 points | 6/13 |
| Stable state of RS | 0.789 (0.750) | -0.7 points | 8/11 |
| Change of stable state | **0.842 (0.798)\*** | -0.2 points | 12/7 |
| **BPI impact score** |  |  |  |
| Dynamic state of RS | 0.684 (0.583) | -0.7 points | 12/7 |
| Stable state of RS | 0.737 (0.682) | -0.8 points | 11/8 |
| Change of stable state | 0.842 (0.731) | -0.1 points | 13/6 |
| **painDETECT intensity score** |  |  |  |
| Dynamic state of RS | **0.944 (0.877)\*** | -2 points | 5/13 |
| Stable state of RS | **0.889 (0.912)\*** | -1 points | 8/10 |
| Change of stable state | 0.778 (0.700) | -1 points | 8/10 |
| **painDETECT total score** |  |  |  |
| Dynamic state of RS | 0.632 (0.557) | -2 points | 8/11 |
| Stable state of RS | 0.579 (0.477) | -2 points | 8/11 |
| Change of stable state | 0.789 (0.486) | -3 points | 5/13 |

### 3.1 Dynamic RS

Classification of patients to those likely or unlikely to benefit from rTMS utilizing the dynamic state RS showed that classification performed best when it was done based on the painDETECT intensity score (Fig. 3A), but also well with the BPI severity score (Table 2): the classification based on painDETECT intensity score indicated that the transition from the dynamic to the stable state, *i.e.* the suppression of MEP amplitude, was significantly stronger in those likely to benefit from the rTMS (*p* = 0.014, Fig. 3B) and corresponding when classification was made based on the BPI severity score (*p* = 0.022). The optimal cut-off points for the dynamic RS were found to be 0.16 and 0.32, respectively. Dynamic state RS did not show predictive power with any other parameter.

**Figure 3:**
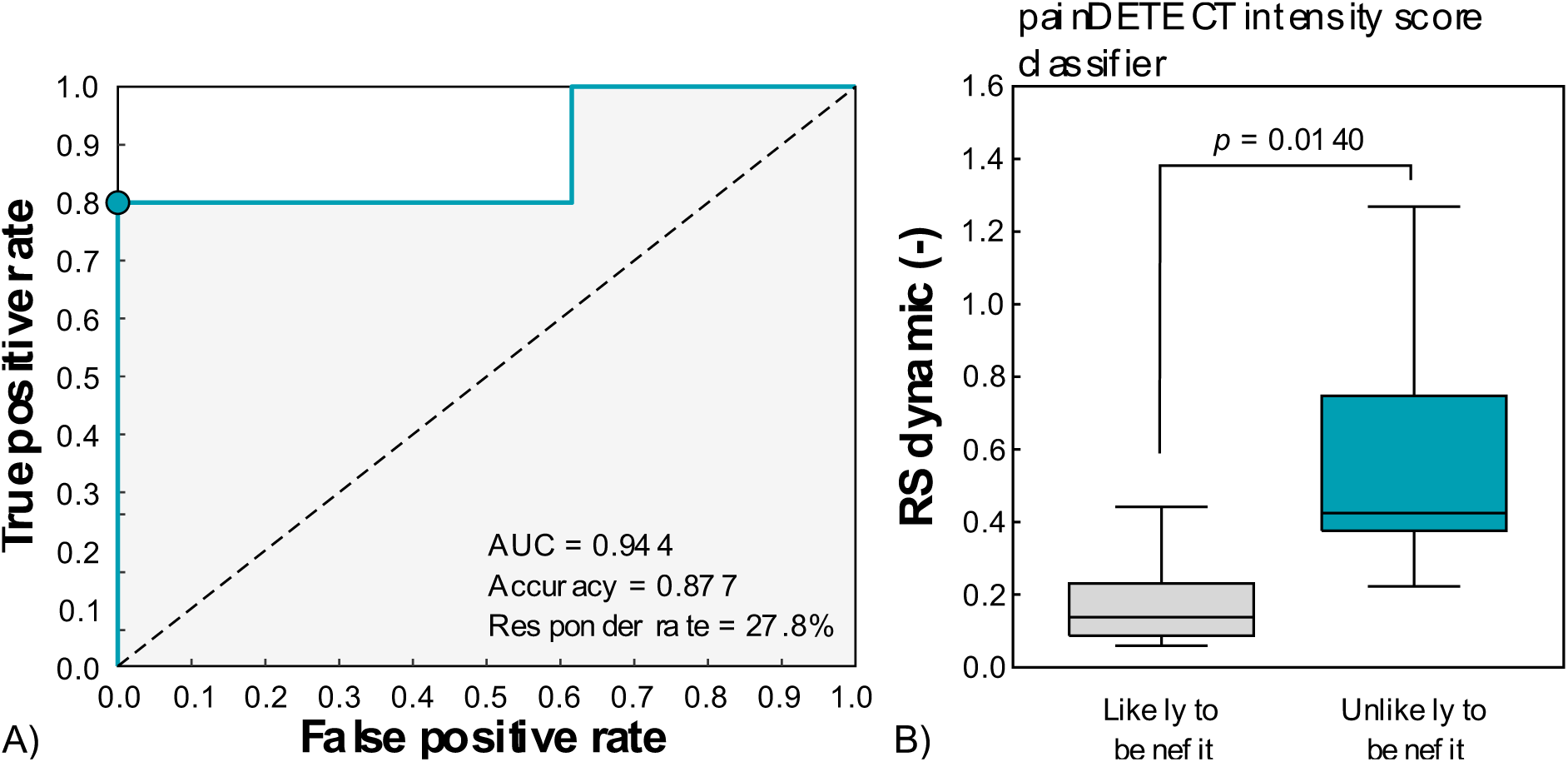
A) Receiving operating characteristic (ROC) curve to identify the patients likely vs. those unlikely to benefit from rTMS based on the dynamic state of the RS. The responders were defined as those showing decrease in painDETECT intensity score in last treatment session. B) MEP amplitude responses in dynamic RS response at baseline level, the two clusters of patients based on the painDETECT intensity score. The suppression of excitability from the dynamic state to the suppressed stable state, *i.e.* the dynamic RS response, was significantly greater in those who displayed analgesic effect. In the plot, the box represents the 25th and 75th quantile with horizontal line representing the median and the whiskers represent the 5th and 95th quantile. The *p*-value indicates the significance of the difference based on Mann-Whitney U test. The (turquoise) dot in figure A exhibits the optimum threshold to achieve the maximum area under the ROC curve (AUC).

### 3.2 Stable RS

When classifying patients to likely or unlikely to benefit by using stable state RS, we found that it performed best when classification was done based on the painDETECT intensity score (Fig. 4A, Table 2). The optimal cut-off point for stable RS was found to be 142µV. The classification indicated that the stable state MEP amplitude level before rTMS therapy was significantly lower in those likely to benefit from the rTMS when classification was made on the basis of painDETECT intensity score (*p* = 0.002, Fig. 4B, Fig. 5A).

**Figure 4.**
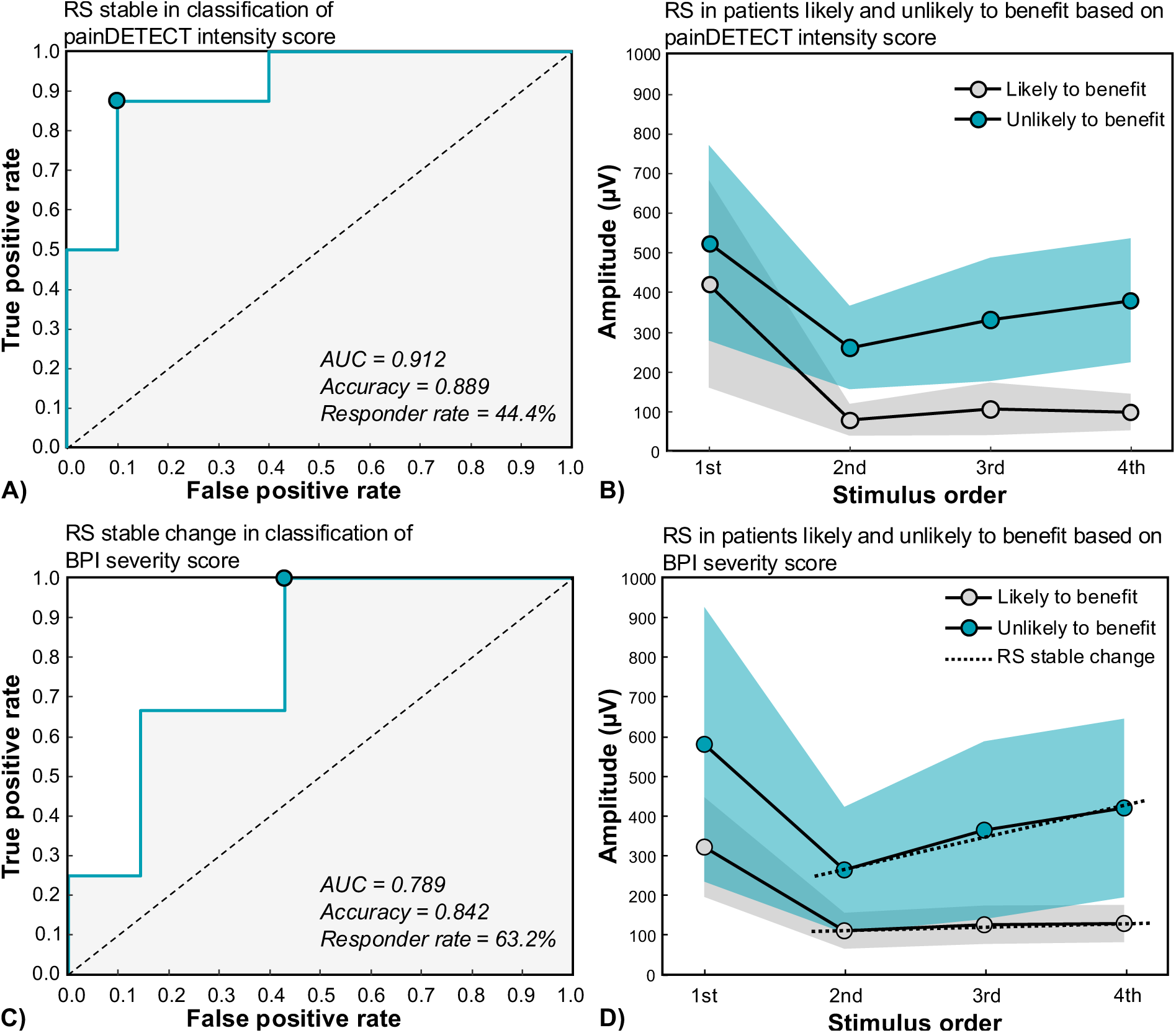
A) Receiving operating characteristic (ROC) curve to identify the patients likely vs. those unlikely to benefit from rTMS based on the stable state of the RS. The responders were defined as those showing decrease in painDETECT intensity score in last treatment session. B) MEP amplitude (mean ± standard error of RS, within each cluster of patients before the initial rTMS session based on the painDETECT intensity score. C) ROC curve to discriminate the patients showing analgesic effect in response to rTMS from those who did not, based on the change of stable state of RS throughout the second to the fourth evoked responses. Analgesic effect was considered as the decrease in BPI severity score. D) MEP amplitude (mean ± standard error of RS, within each cluster of patients before the initial rTMS session based on the BPI severity score. The turquoise dot in figures A and C exhibit the optimum threshold to achieve the maximum area under the ROC curve (AUC).

**Figure 5.**
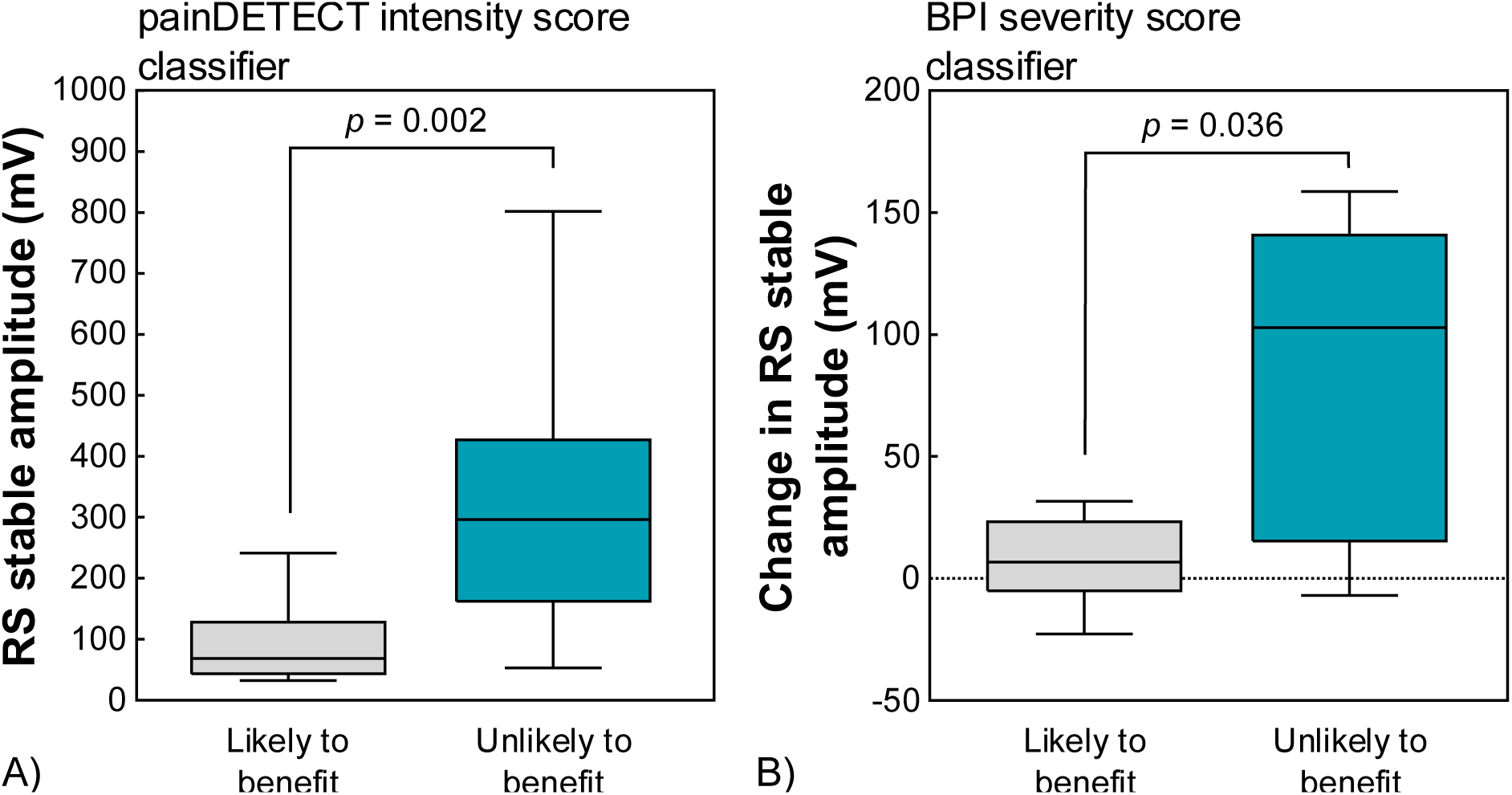
A) MEP amplitude responses in stable RS at baseline level, the two clusters of patients based on the painDETECT intensity score. The overall excitability, evident through the mean MEP amplitude over the suppressed responses, was significantly lower in those who showed analgesic effect and improvement in quality of life. This might reflect the unsaturated inhibitory tone which allows the rTMS to exert its effect. B) MEP amplitude responses of changes in stable RS at baseline level, between the patients who benefited and did not benefit from rTMS based on change in BPI severity score. The change in the amplitude of the suppressed responses was significantly lower in patients who reported benefit. In the plots, the box represents the 25th and 75th quantile with horizontal line representing the median and the whiskers represent the 5th and 95th quantile. The *p*-values indicate the significance of the difference based on Mann-Whitney U test.

The changes in stable state of the RS in identifying patients likely to experience analgesic effect based on the BPI severity score displayed promising accuracy (Table 2, Fig. 4C) with the optimal cut-off point of 94µV. The classification based on the BPI severity score indicated that the change in stable state MEP amplitude level observed during RS before rTMS therapy was significantly lower in those likely to benefit from the rTMS (*p* = 0.036) (Fig. 4D, Fig. 5B). RS stable or change in RS stable did not display significant effects with any other parameter.

## 4 Discussion

The findings from the present study suggest the potential predictive value of RS for rTMS-induced analgesic effects. For immediate subjective responsiveness to rTMS treatment, the BPI severity score and the painDETECT intensity score appeared as most effective scores in the evaluation of RS’s predictive potential. It has been suggested that the changes in pain intensity, which is addressed in most questionnaires, may not be sufficient in determining the analgesic effect. Hence, we included the scores on interference-items in BPI questionnaire to identify patients who experience analgesic effect (Ballantyne & Sullivan, 2015; Vartiainen et al., 2016). However, none of the scores used in the present study for testing other aspects of pain than the intensity demonstrated any classification potential in connection to RS. One reason may be that the questionnaires were filled just before the treatment and right after it, and the impact of therapy could take longer to arise. It may also be variable in the current study population as the treatment periods varied between one and two weeks and not all patients were naïve to the treatment. There were no significant differences in the outcome scores between those who received one or two weeks of treatment (*p*>0.198).

The TMS-induced RS paradigm in the current study implies the interplay of two states of the neural system, to promote the concurrent observed adaptability (dynamic state) and stability (stable state). The stable state itself could be viewed from two separate yet interwoven aspects; one depicting the overall suppressed excitability that is less affected by the ongoing oscillatory brain dynamics and one showing the recovery of the evoked responses after exposure to the repeated stimuli. Investigating these two aspects, we demonstrated significantly higher overall neural activity and faster recovery in patients reporting no benefit of treating their chronic pain with rTMS. Multiple lines of evidence suggest that the chronic pain is associated with anatomical (volume loss) and biochemical changes (GABA content) of the thalamus, resulting in the altered thalamocortical connectivity and loss of inhibition (Apkarian et al., 2004; Henderson et al., 2013; Hong et al., 2010; Pattany et al., 2002).

Despite conflicting findings, some studies in neuropathic chronic pain have exhibited that this disruption in excitation/inhibition balance has left the motor cortex with a state of disinhibition (Chang et al., 2018; Kirveskari et al., 2010; Lefaucheur et al., 2006). The analgesic effects observed in high-frequency rTMS could be attributed to restoring this balance through unleashing the inhibitory GABA tone at the dorsal-horn (Leung et al., 2009). Hence, the lack of analgesic effect in subpopulation of the patients could be explained via the saturated inhibitory tone, reflected through the significantly higher overall activity at stable level, which does not allow for the 10-Hz rTMS to induce further inhibition. While the GABA-mediated inhibition could be the underpinning mechanism of the first aspect of the RS (Duque et al., 2014; Natan et al., 2015; Perez-Gonzalez et al., 2012), the second aspect, recovery of the TMS-evoked responses (change in stable state), could result from the modifications at synaptic level. The minor changes in the amplitudes of the consecutive suppressed responses might point to the changes in the strength and efficacy of synapses (Kariminezhad et al., 2020). The employed short-term synaptic plasticity could result in the formation of an “automatic memory” trace that can be detected in ITIs shorter than 3 s (Loewenstein et al., 2015; Minerbi et al., 2009; Pitkänen et al., 2017; Susman et al., 2019). In other words, the low variance and minor recovery in the MEP amplitudes within stable RS in patients showing benefit might imply their potential to hold a memory trace from a recently delivered stimulus. This could in turn facilitate the efficient processing of the repeated stimulus, seeking to minimize the free energy, and probably hinder processing it as a novel stimulus (Friston, 2005). Hence, the shortage of this capacity, as evident in patients with no analgesic effect, can result in the fast recovery of the second and later evoked responses, attributable to the disrupted homeostatic plasticity. Although the Hebbian synaptic plasticity provides the substrate for the memory (Bliss & Collingridge, 1993; Voronin, 1983), the synaptic homeostasis is the backbone of the functional outputs, without which the brain would succumb to either hyper- or hypo-activity resulting in pathological states (Litwin-Kumar & Doiron, 2014; Tetzlaff et al., 2011; Zenke et al., 2015). Evidence of such positive feedback instability arising from the unidirectional modification of the synaptic efficacy has been observed in hyperalgesia, where a mildly noxious stimulus causes disproportionate pain (Baller & Ross, 2017; Basbaum et al., 2009). Also, impaired homeostatic plasticity in the M1 has been proposed as the plausible pathophysiological mechanism for chronic low back pain (Thapa et al., 2018). Taking these into account and as synaptic plasticity has been assumed as one key mechanism underlying the therapeutic effect of rTMS (Chervyakov et al., 2015; Hoogendam et al., 2010), assessing the change in the stable state of the RS can provide the information on the capacity of the synaptic efficacy to undergo the required modification following neuromodulation therapies.

An alternative perspective on the sustained salience experienced in response to the repeated stimulus suggests the involvement of the emotional factor of the chronic pain along with the sensory one (Borsook et al., 2013; Colloca et al., 2004; Levine & Gordon, 1984). Examples supporting this notion include studies in which pain was perceived in the absence of sensory painful stimuli, or pain relief reported in the absence of an analgesic medication (Danziger et al., 2009; Hashmi et al., 2012). These findings provide significant insights into the role of the baseline innate state of the brain in assigning salience values to the sensory stimuli. Hence, the aberrant network underpinning salience processing may partially account for the sustained salience and deficits in adaptation as observed. Main regions that have been suggested to be involved in this network are the basal ganglia and the thalamus, the regions through which the RS is also mediated (Borsook et al., 2013; Julkunen et al., 2018).

Taking the aforementioned aspects, as RS at stable level can plausibly capture both the overall excitability due to *e.g.* the changes in GABA content, and the modifications in evoked suppressed responses, due to *e.g.* the changes in homeostatic synaptic plasticity, this can introduce TMS-evoked RS as an objective predictive biomarker which complements the widely-used subjective assessment. While the subjective assessment of pain cannot be disregarded, it alone may not provide early sufficient data for therapeutic decision-making.

The involvement of emotional and cognitive factors in parallel with lack of analgesic effects in many patients affect the variability due to subjective ratings (Minerbi et al., 2009; Moruzzi & Magoun, 1949).

Some limitations need to be acknowledged in the present study. 1) Given the complexity of the pain as a subjective sensation, influenced by multiple factors, a clinical definition for a “responder” can be ill-defined through questionnaire. The changes in subjective pain rating can be counted on only where the interventions have produced considerable analgesic effect, *e.g.* 30% reduction in VAS scores (Andre-Obadia et al., 2018). However, this definition can disregard those patients experiencing the less but still meaningful improvement in their quality of life (Säisänen et al., 2022). For comparison, when normalizing the change in painDETECT intensity scores with the baseline score, 30% reduction in NRS was experienced by 5/13 patients. This division resulted in accuracy of 0.944 (AUC = 0.846) with significantly (*p*=0.026) greater suppression in the MEPs assessed by dynamic state RS. 2) The small sample size has led us to identify those who benefited from rTMS based on statistical performance. Hence, the classification to those likely/not likely to benefit was made via optimization of outcome scores and differentiation based on the RS responses to identify responsiveness to rTMS prior to the interventions. 3) The target area of treatment was switched from M1 to S2 in seven patients after 5 sessions. This might have had effect on the responsiveness to rTMS; however, as mentioned above the crucial question in this study was regarding the predictive power of RS addressed in M1. 4) No follow-up was made to assess the long-term effect of the intervention where the patients might have experienced late analgesic effect and could have displayed clear changes in the impact of pain on their lives. And finally, 5) although the ROC-analysis suggests the predictive value of RS as a biomarker at individual level, a larger sample size needs to be recruited to assess its applicability for generalization in patient population.

## 5 Conclusions

In conclusion, our results suggest that there is potential in using RS as a predictive marker for identifying patients that could benefit from rTMS therapy for chronic pain, which was observed as analgesic effect immediately after the treatment period in the present study.

Identification of patients who could benefit from rTMS therapy would enable targeting patients with most appropriate means of treatment to optimize outcome, efficiency, and resources.

## Data Availability

Generic data are made available upon personal request for non-commercial use.

## Data availability statement

Generic data are made available upon personal request for non-commercial use.

## Ethics approval statement

The study was approved by the research ethics committee of the Kuopio University Hospital (949/2018).

## Patient consent statement

All patients provided a written informed consent prior to their participation.

## References

Andre-Obadia, N., Magnin, M., Simon, E., & Garcia-Larrea, L. (2018). Somatotopic effects of rTMS in neuropathic pain? A comparison between stimulation over hand and face motor areas. Eur J Pain, 22(4), 707–715. doi:10.1002/ejp.1156

Apkarian, A. V., Bushnell, M. C., Treede, R. D., & Zubieta, J. K. (2005). Human brain mechanisms of pain perception and regulation in health and disease. Eur J Pain, 9(4), 463–484. doi:10.1016/j.ejpain.2004.11.001

Apkarian, A. V., Sosa, Y., Sonty, S., Levy, R. M., Harden, R. N., Parrish, T. B., & Gitelman, D. R. (2004). Chronic back pain is associated with decreased prefrontal and thalamic gray matter density. J Neurosci, 24(46), 10410–10415. doi:10.1523/JNEUROSCI.2541-04.2004

Awiszus, F., & Borckardt, J. (2012). TMS Motor Threshold Assessment Tool 2.0 Retrieved from http://clinicalresearcher.org/software.htm

Ballantyne, J. C., & Sullivan, M. D. (2015). Intensity of Chronic Pain--The Wrong Metric? N Engl J Med, 373(22), 2098–2099. doi:10.1056/NEJMp1507136

Baller, E. B., & Ross, D. A. (2017). Your System Has Been Hijacked: The Neurobiology of Chronic Pain. Biol Psychiatry, 82(8), e61–e63. doi:10.1016/j.biopsych.2017.08.009

Basbaum, A. I., Bautista, D. M., Scherrer, G., & Julius, D. (2009). Cellular and molecular mechanisms of pain. Cell, 139(2), 267–284. doi:10.1016/j.cell.2009.09.028

Bavelier, D., & Neville, H. J. (2002). Cross-modal plasticity: where and how? Nat Rev Neurosci, 3(6), 443–452. doi:10.1038/nrn848

Bliss, T. V., & Collingridge, G. L. (1993). A synaptic model of memory: long-term potentiation in the hippocampus. Nature, 361(6407), 31–39. doi:10.1038/361031a0

Borsook, D., Edwards, R., Elman, I., Becerra, L., & Levine, J. (2013). Pain and analgesia: the value of salience circuits. Prog Neurobiol, 104, 93–105. doi:10.1016/j.pneurobio.2013.02.003

Brown, A., & Weaver, L. C. (2012). The dark side of neuroplasticity. Exp Neurol, 235(1), 133–141. doi:10.1016/j.expneurol.2011.11.004

Cappelleri, J. C., Bienen, E. J., Koduru, V., & Sadosky, A. (2014). Measurement properties of painDETECT by average pain severity. Clinicoecon Outcomes Res, 6, 497–504. doi:10.2147/CEOR.S68997

Chang, W. J., O’Connell, N. E., Beckenkamp, P. R., Alhassani, G., Liston, M. B., & Schabrun, S. M. (2018). Altered Primary Motor Cortex Structure, Organization, and Function in Chronic Pain: A Systematic Review and Meta-Analysis. J Pain, 19(4), 341–359. doi:10.1016/j.jpain.2017.10.007

Chervyakov, A. V., Chernyavsky, A. Y., Sinitsyn, D. O., & Piradov, M. A. (2015). Possible Mechanisms Underlying the Therapeutic Effects of Transcranial Magnetic Stimulation. Front Hum Neurosci, 9, 303. doi:10.3389/fnhum.2015.00303

Cleeland, C. S., & Ryan, K. M. (1994). Pain assessment: global use of the Brief Pain Inventory. Ann Acad Med Singapore, 23, 129–138.

Colloca, L., Lopiano, L., Lanotte, M., & Benedetti, F. (2004). Overt versus covert treatment for pain, anxiety, and Parkinson’s disease. Lancet Neurol, 3(11), 679–684. doi:10.1016/S1474-4422(04)00908-1

Cruccu, G., Garcia-Larrea, L., Hansson, P., Keindl, M., Lefaucheur, J. P., Paulus, W., . . . Attal, N. (2016). EAN guidelines on central neurostimulation therapy in chronic pain conditions. Eur J Neurol, 23(10), 1489–1499. doi:10.1111/ene.13103

Dansie, E. J., & Turk, D. C. (2013). Assessment of patients with chronic pain. Br J Anaesth, 111(1), 19–25. doi:10.1093/bja/aet124

Danziger, N., Faillenot, I., & Peyron, R. (2009). Can we share a pain we never felt? Neural correlates of empathy in patients with congenital insensitivity to pain. Neuron, 61(2), 203–212. doi:10.1016/j.neuron.2008.11.023

Daut, R. L., Cleeland, C. S., & Flanery, R. C. (1983). Development of the Wisconsin Brief Pain Questionnaire to assess pain in cancer and other diseases. Pain, 17(2), 197–210. doi:10.1016/0304-3959(83)90143-4

Duque, D., Malmierca, M. S., & Caspary, D. M. (2014). Modulation of stimulus-specific adaptation by GABA(A) receptor activation or blockade in the medial geniculate body of the anaesthetized rat. J Physiol, 592(4), 729–743. doi:10.1113/jphysiol.2013.261941

Finnerup, N. B., Sindrup, S. H., & Jensen, T. S. (2010). The evidence for pharmacological treatment of neuropathic pain. Pain, 150(3), 573–581. doi:10.1016/j.pain.2010.06.019

Freynhagen, R., Baron, R., Gockel, U., & Tolle, T. R. (2006). painDETECT: a new screening questionnaire to identify neuropathic components in patients with back pain. Curr Med Res Opin, 22(10), 1911–1920. doi:10.1185/030079906X132488

Friston, K. (2005). A theory of cortical responses. Philos Trans R Soc Lond B Biol Sci, 360(1456), 815–836. doi:10.1098/rstb.2005.1622

Grill-Spector, K., Henson, R., & Martin, A. (2006). Repetition and the brain: neural models of stimulus-specific effects. Trends Cogn Sci, 10(1), 14–23. doi:10.1016/j.tics.2005.11.006

Hashmi, J. A., Baria, A. T., Baliki, M. N., Huang, L., Schnitzer, T. J., & Apkarian, V. A. (2012). Brain networks predicting placebo analgesia in a clinical trial for chronic back pain. Pain, 153(12), 2393–2402. doi:10.1016/j.pain.2012.08.008

Henderson, L. A., Peck, C. C., Petersen, E. T., Rae, C. D., Youssef, A. M., Reeves, J. M., . . . Gustin, S. M. (2013). Chronic pain: lost inhibition? J Neurosci, 33(17), 7574–7582. doi:10.1523/JNEUROSCI.0174-13.2013

Hong, J. H., Bai, D. S., Jeong, J. Y., Choi, B. Y., Chang, C. H., Kim, S. H., Jang, S. H. (2010). Injury of the spino-thalamo-cortical pathway is necessary for central post-stroke pain. Eur Neurol, 64(3), 163–168. doi:10.1159/000319040

Hoogendam, J. M., Ramakers, G. M., & Di Lazzaro, V. (2010). Physiology of repetitive transcranial magnetic stimulation of the human brain. Brain Stimul, 3(2), 95–118. doi:10.1016/j.brs.2009.10.005

Julkunen, P., Löfberg, O., Kallioniemi, E., Hyppönen, J., Kälviäinen, R., & Mervaala, E. (2018). Abnormal motor cortical adaptation to external stimulus in Unverricht-Lundborg disease (progressive myoclonus type 1, EPM1). J Neurophysiol, 120(2), 617–623. doi:10.1152/jn.00063.2018

Julkunen, P., Säisänen, L., Danner, N., Niskanen, E., Hukkanen, T., Mervaala, E., & Könönen, M. (2009). Comparison of navigated and non-navigated transcranial magnetic stimulation for motor cortex mapping, motor threshold and motor evoked potentials. Neuroimage, 44(3), 790–795. doi:10.1016/j.neuroimage.2008.09.040

Julkunen, P., Säisänen, L., Hukkanen, T., Danner, N., & Könönen, M. (2012). Does second-scale intertrial interval affect motor evoked potentials induced by single-pulse transcranial magnetic stimulation? Brain Stimul, 5(4), 526–532. doi:10.1016/j.brs.2011.07.006

Kallioniemi, E., Savolainen, P., Järnefelt, G., Koskenkorva, P., Karhu, J., & Julkunen, P. (2018). Transcranial magnetic stimulation modulation of corticospinal excitability by targeting cortical I-waves with biphasic paired-pulses. Brain Stimul, 11(2), 322–326. doi:10.1016/j.brs.2017.10.014

Kariminezhad, S., Karhu, J., Säisänen, L., Reijonen, J., Könönen, M., & Julkunen, P. (2020). Brain Response Induced with Paired Associative Stimulation Is Related to Repetition Suppression of Motor Evoked Potential. Brain Sci, 10(10). doi:10.3390/brainsci10100674

Kirveskari, E., Vartiainen, N. V., Gockel, M., & Forss, N. (2010). Motor cortex dysfunction in complex regional pain syndrome. Clin Neurophysiol, 121(7), 1085–1091. doi:10.1016/j.clinph.2010.01.032

Lefaucheur, J. P. (2006). The use of repetitive transcranial magnetic stimulation (rTMS) in chronic neuropathic pain. Neurophysiol Clin, 36(3), 117–124. doi:10.1016/j.neucli.2006.08.002

Lefaucheur, J. P., Aleman, A., Baeken, C., Benninger, D. H., Brunelin, J., Di Lazzaro, V., . . . Ziemann, U. (2020). Evidence-based guidelines on the therapeutic use of repetitive transcranial magnetic stimulation (rTMS): An update (2014-2018). Clin Neurophysiol, 131(2), 474–528. doi:10.1016/j.clinph.2019.11.002

Lefaucheur, J. P., Drouot, X., Menard-Lefaucheur, I., Keravel, Y., & Nguyen, J. P. (2006). Motor cortex rTMS restores defective intracortical inhibition in chronic neuropathic pain. Neurology, 67(9), 1568–1574. doi:10.1212/01.wnl.0000242731.10074.3c

Lefaucheur, J. P., Drouot, X., & Nguyen, J. P. (2001). Interventional neurophysiology for pain control: duration of pain relief following repetitive transcranial magnetic stimulation of the motor cortex. Neurophysiol Clin, 31(4), 247–252. doi:10.1016/s0987-7053(01)00260-x

Leung, A., Donohue, M., Xu, R., Lee, R., Lefaucheur, J. P., Khedr, E. M., . . . Chen, R. (2009). rTMS for suppressing neuropathic pain: a meta-analysis. J Pain, 10(12), 1205–1216. doi:10.1016/j.jpain.2009.03.010

Levine, J. D., & Gordon, N. C. (1984). Influence of the method of drug administration on analgesic response. Nature, 312(5996), 755–756. doi:10.1038/312755a0

Litwin-Kumar, A., & Doiron, B. (2014). Formation and maintenance of neuronal assemblies through synaptic plasticity. Nat Commun, 5, 5319. doi:10.1038/ncomms6319

Loewenstein, Y., Yanover, U., & Rumpel, S. (2015). Predicting the Dynamics of Network Connectivity in the Neocortex. J Neurosci, 35(36), 12535–12544. doi:10.1523/JNEUROSCI.2917-14.2015

Löfberg, O., Julkunen, P., Pääkkönen, A., & Karhu, J. (2014). The auditory-evoked arousal modulates motor cortex excitability. Neuroscience, 274, 403–408. doi:10.1016/j.neuroscience.2014.05.060

Löfberg, O., Julkunen, P., Tiihonen, P., Pääkkönen, A., & Karhu, J. (2013). Repetition suppression in the cortical motor and auditory systems resemble each other--a combined TMS and evoked potential study. Neuroscience, 243, 40–45. doi:10.1016/j.neuroscience.2013.03.060

Minerbi, A., Kahana, R., Goldfeld, L., Kaufman, M., Marom, S., & Ziv, N. E. (2009). Long-term relationships between synaptic tenacity, synaptic remodeling, and network activity. PLoS Biol, 7(6), e1000136. doi:10.1371/journal.pbio.1000136

Moruzzi, G., & Magoun, H. W. (1949). Brain stem reticular formation and activation of the EEG. Electroencephalogr Clin Neurophysiol, 1(4), 455–473.

Natan, R. G., Briguglio, J. J., Mwilambwe-Tshilobo, L., Jones, S. I., Aizenberg, M., Goldberg, E. M., & Geffen, M. N. (2015). Complementary control of sensory adaptation by two types of cortical interneurons. Elife, 4. doi:10.7554/eLife.09868

O’Connell, N. E., Marston, L., Spencer, S., DeSouza, L. H., & Wand, B. M. (2018). Non-invasive brain stimulation techniques for chronic pain. Cochrane Database Syst Rev, 4, CD008208. doi:10.1002/14651858.CD008208.pub5

Pascual-Leone, A., Amedi, A., Fregni, F., & Merabet, L. B. (2005). The plastic human brain cortex. Annu Rev Neurosci, 28, 377–401. doi:10.1146/annurev.neuro.27.070203.144216

Pattany, P. M., Yezierski, R. P., Widerstrom-Noga, E. G., Bowen, B. C., Martinez-Arizala, A., Garcia, B. R., & Quencer, R. M. (2002). Proton magnetic resonance spectroscopy of the thalamus in patients with chronic neuropathic pain after spinal cord injury. AJNR Am J Neuroradiol, 23(6), 901–905.

Perez-Gonzalez, D., Hernandez, O., Covey, E., & Malmierca, M. S. (2012). GABA(A)- mediated inhibition modulates stimulus-specific adaptation in the inferior colliculus. PLoS One, 7(3), e34297. doi:10.1371/journal.pone.0034297

Pitkänen, M., Kallioniemi, E., & Julkunen, P. (2017). Effect of inter-train interval on the induction of repetition suppression of motor-evoked potentials using transcranial magnetic stimulation. PLoS One, 12(7), e0181663. doi:10.1371/journal.pone.0181663

Susman, L., Brenner, N., & Barak, O. (2019). Stable memory with unstable synapses. Nat Commun, 10(1), 4441. doi:10.1038/s41467-019-12306-2

Säisänen, L., Huttunen, J., Hyppönen, J., Nissen, M., Kotiranta, U., Mervaala, E., & Fraunberg, M. (2022). Efficacy and tolerability in patients with chronic facial pain of two consecutive treatment periods of rTMS applied over the facial motor cortex, using protocols differing in stimulation frequency, duration, and train pattern. Neurophysiol Clin, 52(2), 95–108. doi:10.1016/j.neucli.2022.03.001

Tan, G., Jensen, M. P., Thornby, J. I., & Shanti, B. F. (2004). Validation of the Brief Pain Inventory for chronic nonmalignant pain. J Pain, 5(2), 133–137. doi:10.1016/j.jpain.2003.12.005

Tetzlaff, C., Kolodziejski, C., Timme, M., & Worgotter, F. (2011). Synaptic scaling in combination with many generic plasticity mechanisms stabilizes circuit connectivity. Front Comput Neurosci, 5, 47. doi:10.3389/fncom.2011.00047

Thapa, T., Graven-Nielsen, T., Chipchase, L. S., & Schabrun, S. M. (2018). Disruption of cortical synaptic homeostasis in individuals with chronic low back pain. Clin Neurophysiol, 129(5), 1090–1096. doi:10.1016/j.clinph.2018.01.060

Treede, R. D., Rief, W., Barke, A., Aziz, Q., Bennett, M. I., Benoliel, R., . . . Wang, S. J. (2015). A classification of chronic pain for ICD-11. Pain, 156(6), 1003–1007. doi:10.1097/j.pain.0000000000000160

Vartiainen, P., Heiskanen, T., Sintonen, H., Roine, R. P., & Kalso, E. (2016). Health-related quality of life and burden of disease in chronic pain measured with the 15D instrument. Pain, 157(10), 2269–2276. doi:10.1097/j.pain.0000000000000641

Voronin, L. L. (1983). Long-term potentiation in the hippocampus. Neuroscience, 10(4), 1051–1069. doi:10.1016/0306-4522(83)90099-4

Wiffen, P. J., Derry, S., Moore, R. A., Aldington, D., Cole, P., Rice, A. S., . . . Kalso, E. A. (2013). Antiepileptic drugs for neuropathic pain and fibromyalgia - an overview of Cochrane reviews. Cochrane Database Syst Rev (11), CD010567. doi:10.1002/14651858.CD010567.pub2

Zenke, F., Agnes, E. J., & Gerstner, W. (2015). Diverse synaptic plasticity mechanisms orchestrated to form and retrieve memories in spiking neural networks. Nat Commun, 6, 6922. doi:10.1038/ncomms7922

